# Intensive Care Unit Capital-Budgeting Workbench: An Open-Source Decision-Support Application for High-Acuity Investment Planning

**DOI:** 10.1101/2025.08.04.25332977

**Authors:** Mahmoud Alwakeel

## Abstract

**Objective:** Capital budgeting in intensive care units (ICUs) demands rapid, high-stakes investment decisions. We developed an open-source ICU Capital Budgeting Workbench that merges a deterministic finance engine with GPT-based natural-language processing (NLP) to streamline planning.

**Materials and Methods:** The web application couples classic valuation metrics (e.g., net present value, internal rate of return, and payback) with an NLP module that converts multi-year free-text scenarios into structured projections. Users describe projects in everyday language; GPT parses these narratives into year-by-year cash flows and discount rates, after which the engine computes financial metrics and produces interactive sensitivity, scenario, and Monte Carlo analyses.

**Results:** In illustrative cases the workbench translated narrative ICU expansion proposals into five and ten-year cash-flow tables and NPV calculations within seconds, eliminating manual spreadsheet construction. Interactive dashboards let users test key assumptions, instantly revealing how occupancy, reimbursement, or inflation shifts influence returns. Compared with traditional ad-hoc spreadsheets, the tool demonstrated marked time savings and consistent analytic structure.

**Discussion:** This proof-of-concept shows how large language models can reduce transcription errors, standardize methodology, and embed uncertainty modeling in routine capital planning. Limitations include dependence on GPT’s parsing accuracy and the need for real-world validation with authentic hospital data.

**Conclusion:** The ICU Capital Budgeting Workbench exemplifies practical AI integration for finance and operations leaders, offering transparent, reproducible, and scalable decision support for ICU equipment and facility investments. By replacing bespoke spreadsheets with a governed, open-source platform, it may improve efficiency and support better-informed, data-driven investment strategy.

## Introduction

Capital budgeting is a critical process in healthcare organizations for planning major expenditures on technologies, equipment, and infrastructure.[1] In an ICU setting, capital projects such as purchasing ventilators, monitoring systems, or expanding ICU bed capacity often involve multi-million-dollar investments and must be carefully evaluated to ensure that benefits outweigh costs.[2] Standard financial methods like discounted cash flow (DCF) analysis and net present value (NPV) calculations are recommended to assess whether a project’s returns justify its upfront cost.[3] These analyses consider the time value of money by projecting annual cash flows (e.g. increased revenue or cost savings) and discounting them to present value, yielding metrics like NPV and internal rate of return (IRR) that inform decision-makers.[3] Aside from pure financial return, hospital capital decisions may also factor in strategic or mission-driven needs (for example, compliance with new regulations or quality improvements), even if financial ROI is modest.[1]

Despite the importance of rigorous analysis, in practice many hospitals still rely on manual spreadsheets and ad hoc methods for capital budgeting. This is problematic because spreadsheets are notoriously error-prone: one study found that over 90% of spreadsheets in a hospital setting contained critical errors impacting the “bottom line” calculations.[4] Such errors and inconsistencies can lead to suboptimal or even harmful investment decisions. Moreover, the capital planning process is time-consuming; analysts must gather data, build financial models for each proposal, and iterate through multiple scenarios.[5] Differences in modeling approaches between analysts or departments further undermine standardization. There is a clear opportunity to improve the efficiency, accuracy, and consistency of ICU capital budgeting through better software tools and decision support systems.

Recent advances in artificial intelligence (AI) offer a potential solution. Large language models (LLMs) such as GPT have shown remarkable ability to interpret unstructured text and perform complex reasoning tasks in various domains. In clinical decision-making, for example, GPT has been able to provide recommendations from free-text patient case descriptions that approach expert-level judgment.[6] In healthcare operations, LLMs are being explored as assistants to automate and streamline administrative and analytical workflows.[7,8] Hospitals generate vast amounts of textual plans, reports, and proposals; an LLM that can understand and transform such inputs into structured analyses could greatly accelerate processes like budgeting. Early studies suggest that LLM-driven automation can save time and reduce costs for health systems, if deployed with proper oversight.[9]

Within this context, we developed the ICU Capital Budgeting Workbench, an AI-assisted application to support ICU capital expenditure planning. This tool uniquely combines a deterministic finance engine (for standard calculations such as NPV, IRR, payback period, etc.) with a GPT based natural language interface that allows users to describe financial scenarios in plain language. By doing so, it addresses two major pain points: (1) reducing manual data entry and formula errors by automating the conversion of narrative project descriptions into structured financial models, and (2) enforcing a consistent analysis framework across all projects, improving standardization. We hypothesize that this approach can increase efficiency (faster turnaround for budget proposals) and improve decision quality (by minimizing spreadsheet errors and ensuring that each project is evaluated with the same rigorous criteria).

## Materials and Methods

### System Architecture and Design

The ICU Capital Budgeting Workbench is implemented as a three-tier web application (Figure 1). At the presentation tier, a Python/Streamlit interface provides six task-oriented pages; New Project, Dashboard, Reports, Risk Tools, Capital Rationing, and Divisions, that mirror the typical capital-planning workflow. Every page renders entirely in the browser, while state is retained on the server through Streamlit’s session object. Users may enter data in three complementary ways: (1) free-text narratives that describe revenues, costs, discount assumptions, and qualitative risks; (2) structured forms that capture year-by-year numeric values with unlimited sub-elements (e.g., ventilator maintenance, nursing overtime, training expenses, etc); and (3) Excel import/export for portfolio-level hand-offs to accounting systems.

**Figure 1.**
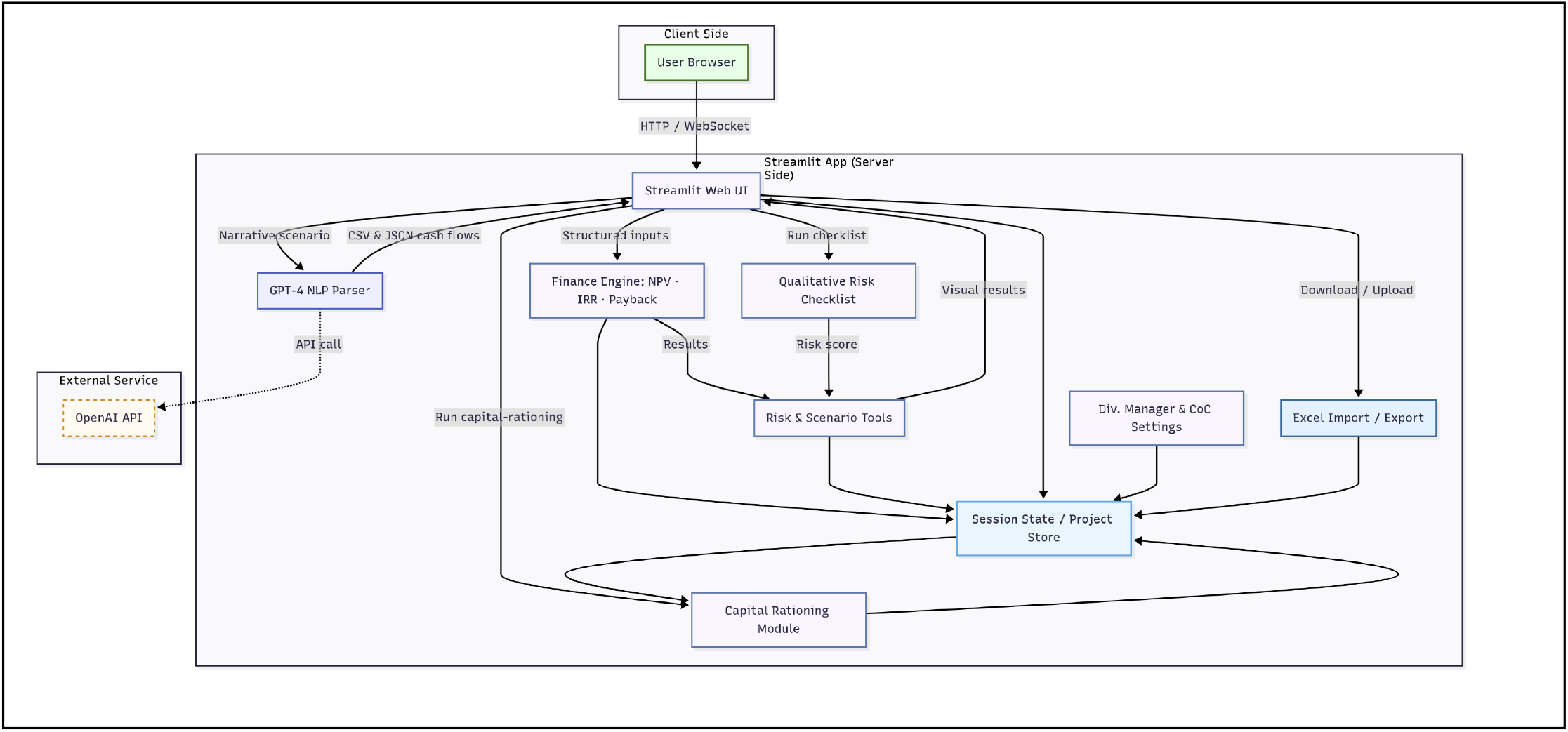
Component diagram of the ICU Capital Budgeting Workbench

The logic tier combines an LLM service with deterministic finance code. A prompt-engineered GPT NLP module exposes two endpoints. The single-item parser converts short phrases such as “maintenance $5,000 per year” into numeric Python expressions, while the multi-year parser transforms longer paragraphs into a three-part payload: a CSV table of annual revenues, costs, and discount rates; a hierarchical JSON dictionary that lists every revenue or cost sub-element; and a narrative explanation that the application displays verbatim so that analysts can inspect, audit, and correct any mis-parsing before calculations proceed (Supplement eFigure 1). Constraining GPT to this rigid schema markedly reduces hallucinations and promotes transparency. Structured outputs are passed to a deterministic finance engine built on numpy_financial python library. For each project the engine computes net-present value (NPV), internal rate of return (IRR), modified IRR, profitability index, and a fractional-year payback period. Parameterization is flexible: users may assign distinct discount rates to individual years and specify a terminal salvage value, enabling faithful modelling of equipment replacements as well as facility expansions.

A surrounding analytics layer implements advanced techniques rarely used in hospital spreadsheets. These include certainty-equivalent valuation, Monte-Carlo simulation, multi-variable sensitivity analysis, and an extended scenario generator that produces worst-, base-, and best-case NPVs together with expected value, standard deviation, coefficient of variation and an automated risk-adjusted discount-rate (RADR) routine. These capabilities are illustrated for the ventilator example in Supplementary eFigures 2-6 (certainty-equivalent valuation, eFigure 2, RADR output, eFigure 3, multi-variable sensitivity, eFigure 4, extended scenario analysis, eFigure 5, Monte-Carlo distribution, eFigure 6). The RADR routine is driven by a four-item Qualitative Risk Checklist captured in the user interface; the checklist asks whether the project involves (i) new or unfamiliar technology, (ii) limited organizational experience, (iii) intense external competition, or (iv) dependence on key personnel. Each affirmative response increments a risk score (0–4), and the RADR module adds 0 pp, 2 pp, or 4 pp to the baseline discount for low-, medium-, or high-risk projects, respectively. The same score is stored with the project to support audit and longitudinal tracking.

Portfolio-level decisions are supported by a capital-rationing page that ranks projects by scenario-specific NPV and highlights when total capital requests exceed the available budget. Administrators may override hurdle rates for individual service lines in the Divisions page; these baseline costs of capital cascade automatically to new projects. All intermediate dictionaries, cash-flow tables, sub-elements, risk scores, analytics outputs, are stored in a session-state object that serves as the data tier; this design eliminates the need for an external database while still allowing complete export to a single Excel workbook for archival review or regulatory audit. Because only rendered tables and plots return to the browser, client overhead remains negligible. The modular design means that any component (e.g., the GPT parser) can be replaced by an on-premises model without refactoring the interface.

### Application workflow

When a user opens New Project, the interface solicits core metadata (project name, category, division), initial investment, useful life, and optional salvage value. The analyst then chooses between manual year-by-year entry or a narrative description. If the narrative option is selected, the paragraph is relayed to GPT; the returned CSV/JSON/explanation bundle is displayed immediately so that the user can verify the parsing, thereby offering an explicit opportunity to detect and correct any LLM error, an important safeguard for financial analyses. Upon acceptance, the deterministic engine runs and appends the computed metrics, cash-flow vectors, sub-element map, and qualitative risk score derived from the checklist to the in-memory portfolio.

The Dashboard lists all projects and shows interactive cash-flow charts; the default line plot (Years 0–N) helps visualize the inflection point at which cumulative net cash flow turns positive (*Figure 2*).

**Figure 2.**
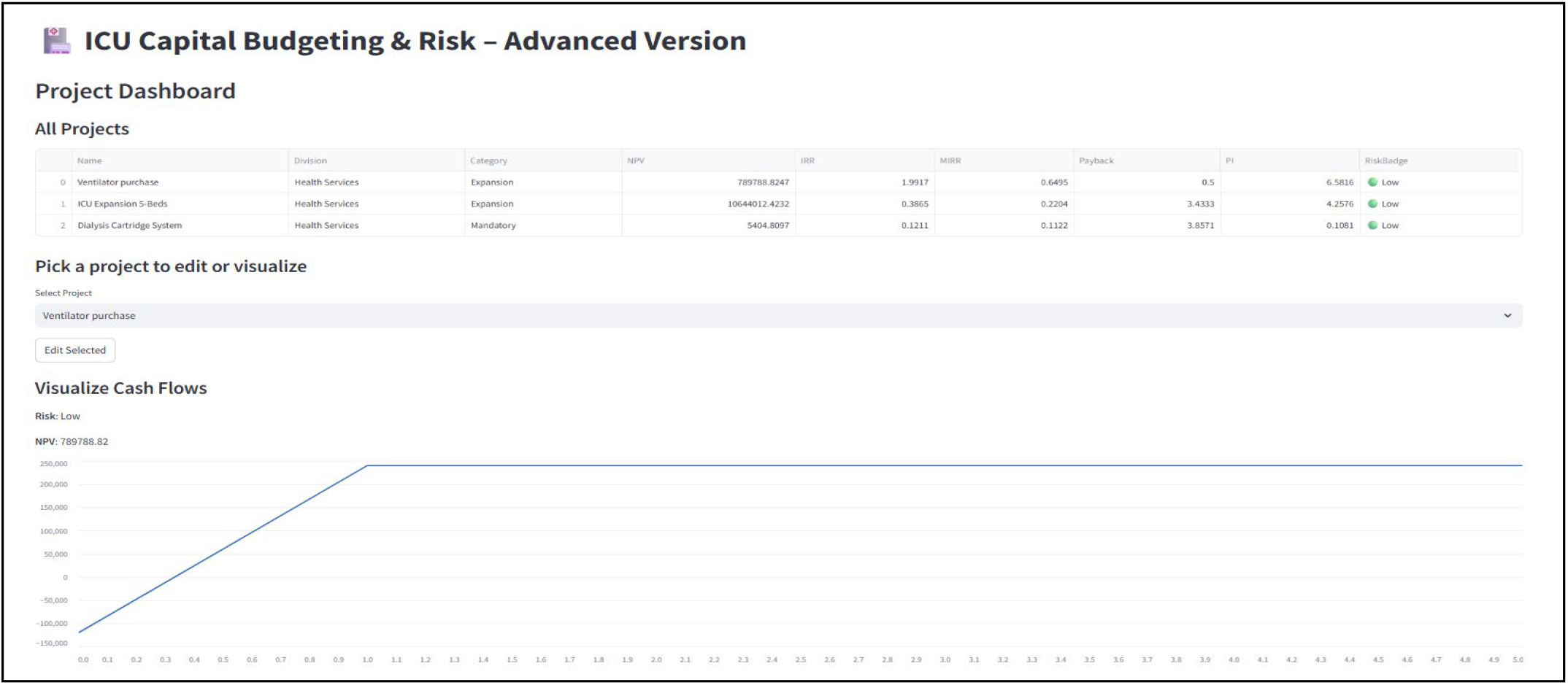
Example cash-flow timeline illustrating the Year 0 capital outlay, subsequent positive net inflows, and the break-even point where cumulative cash flow turns positive.

Selecting a project and opening Risk Tools unlocks the full analytics layer. For example, a user can apply certainty-equivalent factors (0–1) to each year to observe how project value erodes under escalating uncertainty or run a 1,000-trial Monte-Carlo simulation with 20 % volatility in each revenue and cost sub-element to estimate the probability of a negative NPV (Supplement eFigure 2). Outputs are written back to the session store and may later inform the capital-rationing step.

The Reports page generates a portfolio-wide Excel file that contains every cash-flow schedule, sub-element table, metric, and stored risk flag, thereby supporting downstream reconciliation with the hospital’s enterprise resource-planning software as well as retrospective auditing

### Data sources, security, and deployment

Because no live hospital portfolio was available at development time, the demonstration instance uses synthetic scenarios such as ventilator purchases and ICU bed expansions; no protected health information is handled. GPT calls contain only financial text, but the architecture permits substitution with a locally hosted LLM for organizations requiring full data sovereignty. The open-source repository bundles all source code and environment files; a typical calculation-intensive session (one project plus risk analysis) costs only a few US cents in GPT usage and runs comfortably on a standard laptop or containerized server. The application, including source code and deployment instructions, is freely available at https://github.com/Mahmoud-Alwakeel-MD-MMCi/ICU-Capital-Budgeting-Workbench.

### Adaptability and Institutional Tailoring

Although the current demonstration instance targets ICU projects, the workbench was deliberately engineered for rapid localization to any service line or hospital. Because each functional block—user-interface forms, GPT prompt templates, finance routines, risk factors, division-level hurdle rates, and Excel mappings—resides in its own module, organizations can extend or prune features without rewriting core code. For example, a pediatric hospital could inject sub-elements for “family-centered amenities” or “certification fees,” while an academic medical center might add research-grant offsets or faculty effort as revenue streams. Additional qualitative-risk items (e.g., *supply-chain fragility* or *regulatory uncertainty*) can be appended to the checklist with a single JSON entry, and new analytics—such as real-option valuation or Greenhouse-gas payback—can be registered by exposing one function to the finance engine.

Scaling to multiple departments involves only three steps: (i) clone the Divisions page to pre-populate unique baseline costs of capital or strategic weightings; (ii) adjust the GPT prompt to recognize department-specific terminology; and (iii) import historical cash-flow patterns via the Excel interface to seed default assumptions. Because the codebase is less than 2 000 lines and packaged as a Docker container, deployment teams comprising finance analysts, clinical engineers, department administrators, supply-chain managers, strategic-planning staff, and health-system IT can iterate on bespoke functionality in days rather than months, ensuring the tool aligns with local governance processes while retaining a common analytic backbone across the enterprise.

## Results

Three hypotheticals but realistic capital proposals were devised to exercise the workbench. *Scenario 1* (Ventilator Upgrade) assumed purchase of two modern ICU ventilators (capital cost $120,000) generating 100 additional ICU patient-days per year at $1,500 per day, with $30,000 annual operating cost and a five-year horizon at a 10 % discount rate. Scenario 2 (ICU-Bed Expansion) modelled construction of five additional ICU beds (capital cost $2.5 million). Occupancy was staged: two beds filled in Year 1, three in Year 2 and all five from Year 3 onward, producing revenues of $1.2 million, $1.8 million and $3.0 million, respectively, against proportionate staffing costs; project life 10 years, discount rate 8 %. Scenario 3 (Dialysis Cartridge System) posited installation of a point-of-care dialysis unit (capital cost $50,000) yielding $35,000 net savings annually for five years at a 10 % discount.

Table 1 summarizes the deterministic and risk-adjusted outputs. The ventilator purchase, though smallest, delivered the highest efficiency (NPV $0.79 M, PI 6.6, zero probability of loss) and the quickest pay-back (< 1 year). The expansion project produced the greatest absolute NPV (∼$10.6 M) with a favorable PI (4.3) but exhibited wider dispersion under stress tests (worst-case NPV –$2.5 M; best-case $21 M). The dialysis system was only marginally NPV-positive ($5 k), showed the highest coefficient of variation (4.3) and a 34 % Monte-Carlo probability of loss.

**Table 1.**
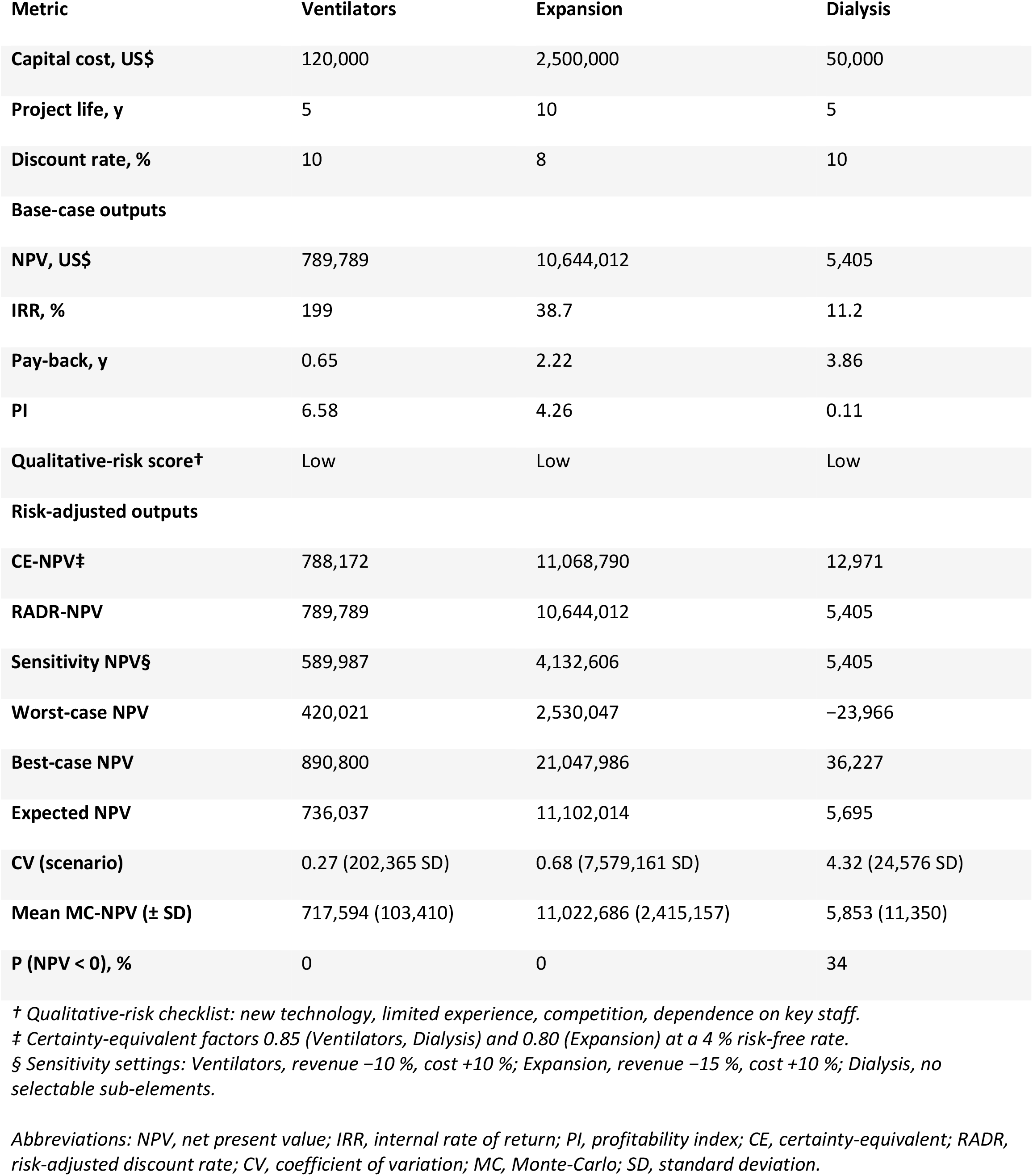
Deterministic and risk-adjusted results for three illustrative ICU projects.

| Metric | Ventilators | Expansion | Dialysis |
| --- | --- | --- | --- |
| Capital cost, US\$ | 120,000 | 2,500,000 | 50,000 |
| Project life, y | 5 | 10 | 5 |
| Discount rate, % | 10 | 8 | 10 |
| **Base-case outputs** |  |  |  |
| NPV, US\$ | 789,789 | 10,644,012 | 5,405 |
| IRR, % | 199 | 38.7 | 11.2 |
| Pay-back, y | 0.65 | 2.22 | 3.86 |
| PI | 6.58 | 4.26 | 0.11 |
| Qualitative-risk score<sup>†</sup> | Low | Low | Low |
| **Risk-adjusted outputs** |  |  |  |
| CE-NPV<sup>‡</sup> | 788,172 | 11,068,790 | 12,971 |
| RADR-NPV | 789,789 | 10,644,012 | 5,405 |
| Sensitivity NPV<sup>§</sup> | 589,987 | 4,132,606 | 5,405 |
| Worst-case NPV | 420,021 | 2,530,047 | −23,966 |
| Best-case NPV | 890,800 | 21,047,986 | 36,227 |
| Expected NPV | 736,037 | 11,102,014 | 5,695 |
| CV (scenario) | 0.27 (202,365 SD) | 0.68 (7,579,161 SD) | 4.32 (24,576 SD) |
| Mean MC-NPV (± SD) | 717,594 (103,410) | 11,022,686 (2,415,157) | 5,853 (11,350) |
| P (NPV < 0), % | 0 | 0 | 34 |

<sup>†</sup> Qualitative-risk checklist: new technology, limited experience, competition, dependence on key staff.
<sup>‡</sup> Certainty-equivalent factors 0.85 (Ventilators, Dialysis) and 0.80 (Expansion) at a 4 % risk-free rate.
<sup>§</sup> Sensitivity settings: Ventilators, revenue −10 %, cost +10 %; Expansion, revenue −15 %, cost +10 %; Dialysis, no selectable sub-elements.
Abbreviations: NPV, net present value; IRR, internal rate of return; PI, profitability index; CE, certainty-equivalent; RADR, risk-adjusted discount rate; CV, coefficient of variation; MC, Monte-Carlo; SD, standard deviation.

From an operational perspective, if capital were severely limited (≤ $0.2 M) the ventilator upgrade would be the sole financially compelling choice (Figure 2). With a mid-sized budget (∼$2.7 M) combining the ventilator and expansion maximizes expected portfolio NPV (> $11 M) while retaining zero simulated downside (Figure 3). The dialysis system should be pursued only if strategic or clinical imperatives outweigh its weak and volatile financial returns or if renegotiated cartridge margins improve its economics appreciably (Supplementary eFigure 19).

The base-case view lists all candidate projects ranked by profitability index. Given the available envelope (black bar, top), the workbench funds the ventilator purchase and dialysis cartridge system (total cost = $170,000) but automatically excludes the five-bed expansion because its cost would breach the limit; the resulting scenario-based portfolio NPV is $0.79 million, and the residual budget is $2.33 million.

**Figure 3.**
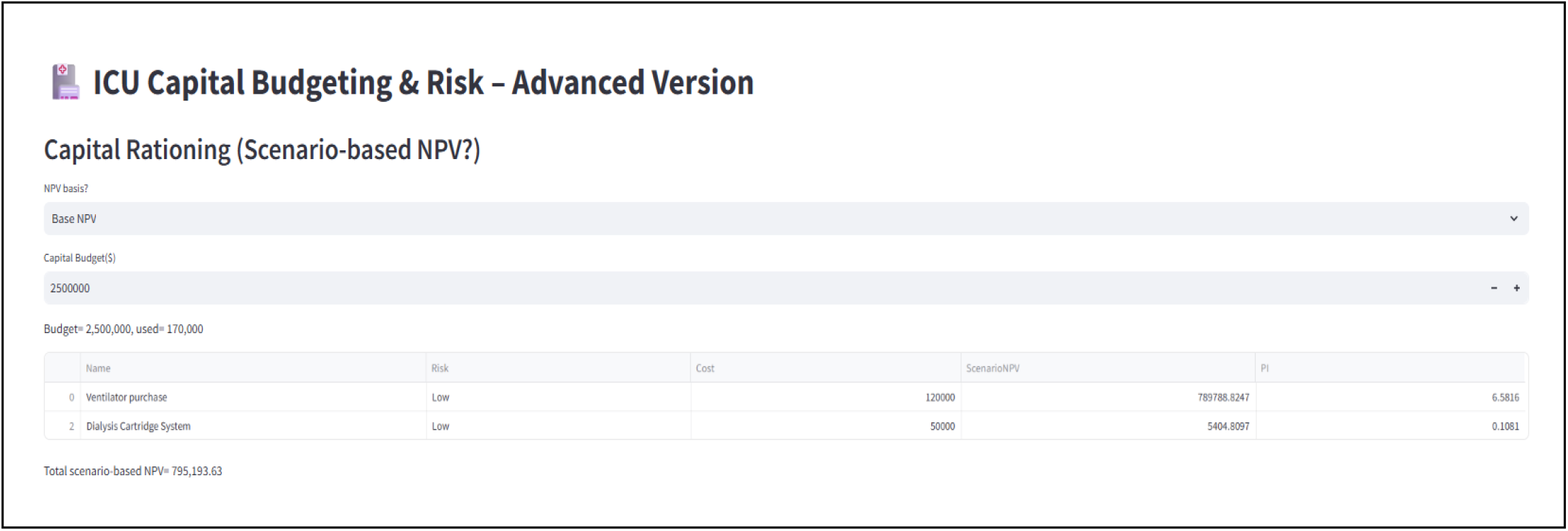
Capital-rationing dashboard under a $2.5 million ICU budget constraint.

## Discussion

The present study introduces and empirically demonstrates an open-source, LLM-enabled workbench that converts free-text ICU capital proposals into auditable, risk-adjusted financial appraisals within minutes. Across three representative scenarios the platform reproduced hand-validated cash-flow models, delivered advanced analytics such as Monte-Carlo simulation, and generated actionable portfolio recommendations—all with no custom coding or spreadsheet manipulation by the user. These findings highlight the tool’s potential to standardize capital-budgeting practice, compress analysis time from hours to seconds, and extend sophisticated risk methods to stakeholders who lack formal finance training. To our knowledge, this is the first study to operationalize a LLM for hospital capital-expenditure planning, distinguishing it from prior LLM work focused primarily on clinical documentation or knowledge retrieval.[6,7]

Robust capital budgeting is a longstanding challenge in hospital operations, especially in critical-care environments where decisions often involve multimillion-dollar equipment or construction commitments.[1] Most U.S. health systems continue to rely on disparate spreadsheet models and manual financial reviews that are susceptible to formula errors and inconsistent assumptions.[4] Commercial capital-planning suites exist, but these platforms focus on enterprise asset tracking and rarely provide fine-grained, project-level risk analytics.[2] Furthermore, they require users to translate clinical narratives into rigid data-entry forms, a step that remains time-consuming and error-prone. The ICU Capital Budgeting Workbench addresses this gap by combining a deterministic finance engine with a LLM parser that automatically converts plain-language business cases into structured cash-flow projections.

Recent literature has documented LLM use in clinical note generation, prior-auth automation, and coding support, with early investigations of LLM in pharmacy inventory modelling.[6–8] However, to our knowledge no prior study has examined an LLM-driven tool for capital-expenditure planning in healthcare. Outside the health sector, LLM has shown competence in corporate-finance tasks such as discounted-cash-flow construction and sensitivity analysis, albeit with performance degradation when prompts are ambiguous.[9] Our experience mirrors these findings. The GPT parser successfully extracted year-by-year revenue, cost, and discount-rate schedules from varied narratives, and its outputs agreed with hand-checked spreadsheets in all illustrative scenarios. Mis-parses were limited to edge cases such as compound sentences with mixed percentage and absolute figures; these were easily identified by reviewing the raw CSV and JSON payloads that the application displays for audit.

The side-by-side results from three hypothetical ICU projects underscore three practical advantages of our approach. First, efficiency: narrative inputs were transformed into full financial models in under one minute, whereas manual spreadsheet construction commonly requires 30 to 60 minutes per project.[10] Second, standardization and accuracy: by embedding vetted algorithms for net-present value, internal rate of return, and risk-adjusted discounting, the tool eliminates silent divergence in formula logic that often arises when analysts maintain personal templates.[4] Third, risk visibility: Monte Carlo simulation, certainty-equivalent valuation, and worst-case scenario analysis are delivered through a single interface, lowering the threshold for finance and clinical leaders to engage with probabilistic metrics. Comparable features are absent from mainstream capital-planning systems and typically require advanced add-ins or separate statistical software.[2,3]

Compared with prior decision-support interventions, the workbench offers a scalable framework that can be tailored to local policy. Any institution-specific element; project categories, hurdle rates, qualitative-risk criteria, or Excel export schema, resides in a configuration file rather than hard-coded logic. A pediatric hospital could, for example, add “family convenience” as a non-financial benefit column or create a separate division with a lower cost of capital to reflect philanthropic subsidies. Because the LLM prompt template is likewise configurable, organizations may substitute department-specific vocabulary (e.g., “negative-pressure room retrofit”) without retraining the model. This modular design supports iterative refinement by finance professionals, biomedical engineers, infection-control leads, and information-technology staff working together on a single code base.

Several limitations warrant caution. The evaluation relied on synthetic data and illustrative scenarios; real-world performance should be assessed in future pilots that measure analyst time savings, error rates, and downstream investment outcomes. The GPT module depends on an external API, raising concerns about cost, latency, and data governance; on-premise or fine-tuned open-weight models are a potential mitigation but require further validation. The current risk-assessment checklist captures four binary factors; more nuanced scales or additional criteria such as supply-chain fragility could improve discrimination. Finally, the workbench focuses on financial return and does not yet integrate non-economic objectives such as equity, environmental sustainability, or alignment with institutional mission. Multi-criteria decision-analysis extensions are technically feasible and would broaden applicability.

## Conclusion

Capital-intensive decisions in intensive-care units have profound clinical and financial consequences. The ICU Capital Budgeting Workbench demonstrates that deterministic finance methods and modern LLMs can be combined to convert unstructured narratives into auditable cash-flow models, deliver advanced risk analytics, and facilitate portfolio optimization within minutes. Illustrative use cases show rapid turn-around, higher standardization, and deeper insight than conventional spreadsheet practice. Although additional field testing is required, the open-source and modular nature of the framework makes it readily adaptable to department-specific needs and evolving organizational criteria. By reducing manual overhead and exposing uncertainty transparently, LLM-augmented capital-budgeting tools have the potential to strengthen fiscal stewardship and, ultimately, to support better-informed investment in critical-care infrastructure.

## Supporting information

eFigure

## Data Availability

The ICU Capital Budgeting Workbench is freely available as open-source software at https://github.com/Mahmoud-Alwakeel-MD-MMCi/ICU-Capital-Budgeting-Workbench

