## Supplementary material for "Intensive Care Unit Capital-Budgeting Workbench: An Open-Source Decision-Support Application for High-Acuity Investment Planning": eFigure

**ICU Capital Budgeting Workbench: GPT-4 Powered Tool for ICU Capital-Expenditure Planning**

**eFigure 1. GPT-4 multi-year parse for the ventilator upgrade (CSV, JSON, explanation)**

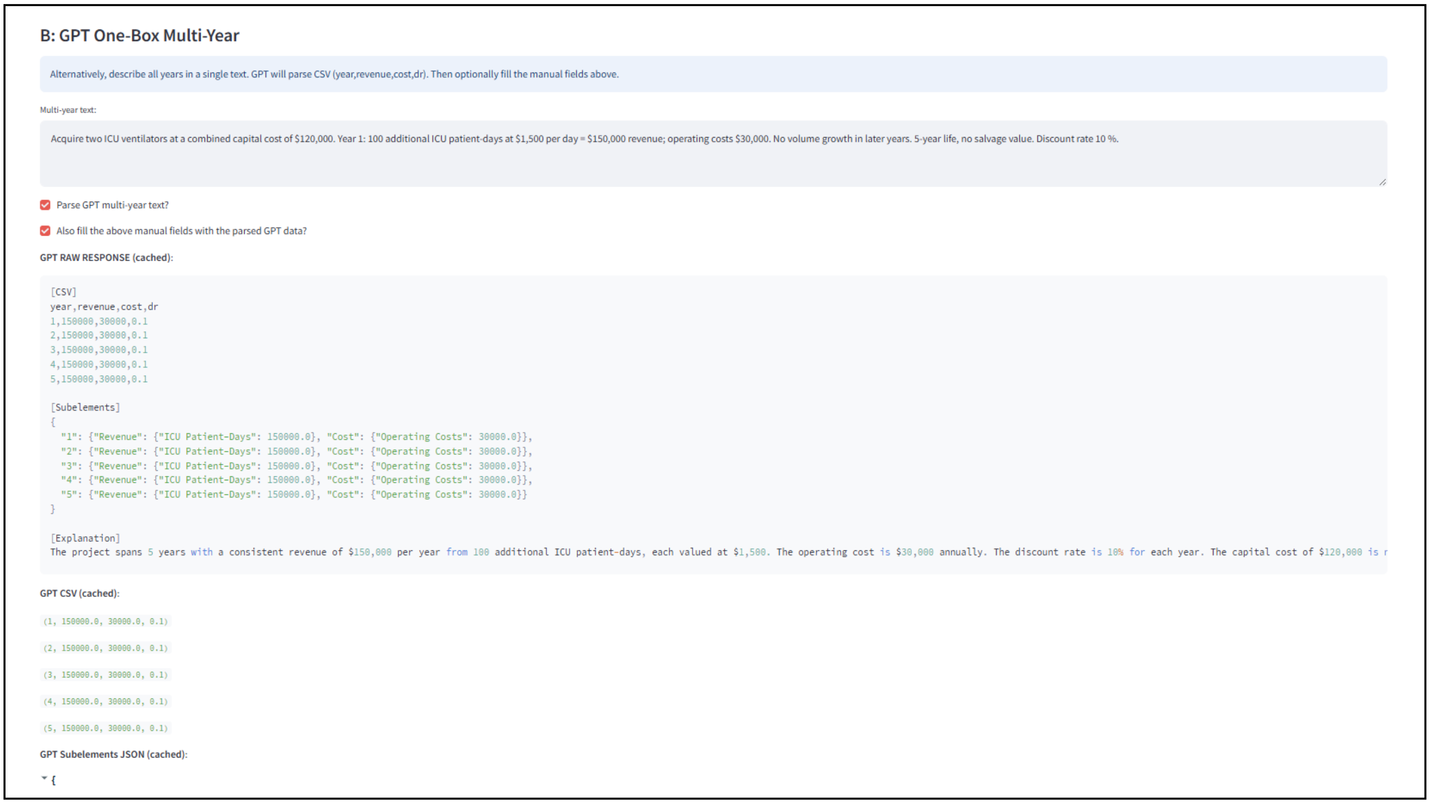

**eFigure 2. Certainty-equivalent sliders and CE-based NPV for the ventilator upgrade.**

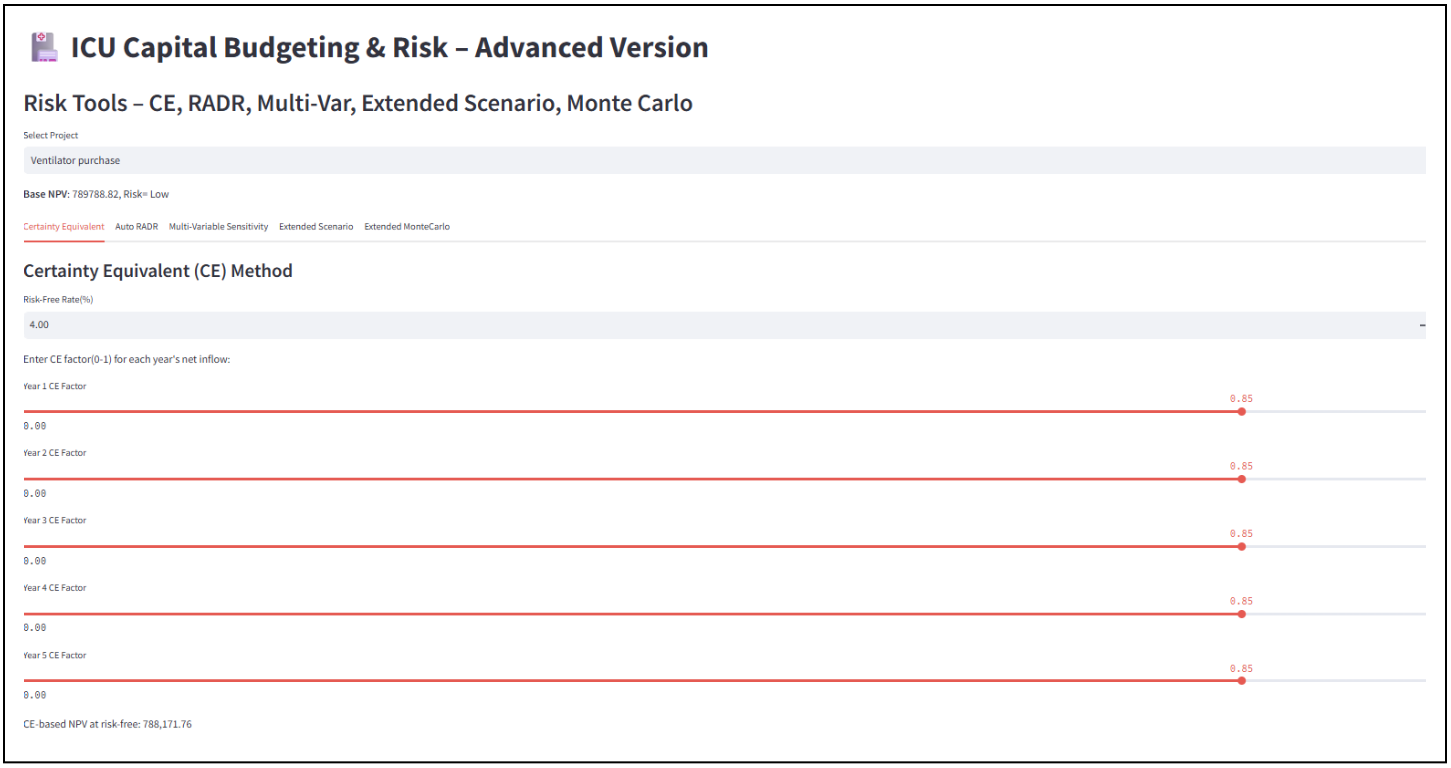

**eFigure 3. Risk-adjusted discount-rate (RADR) output—no adjustment applied (Low risk).**

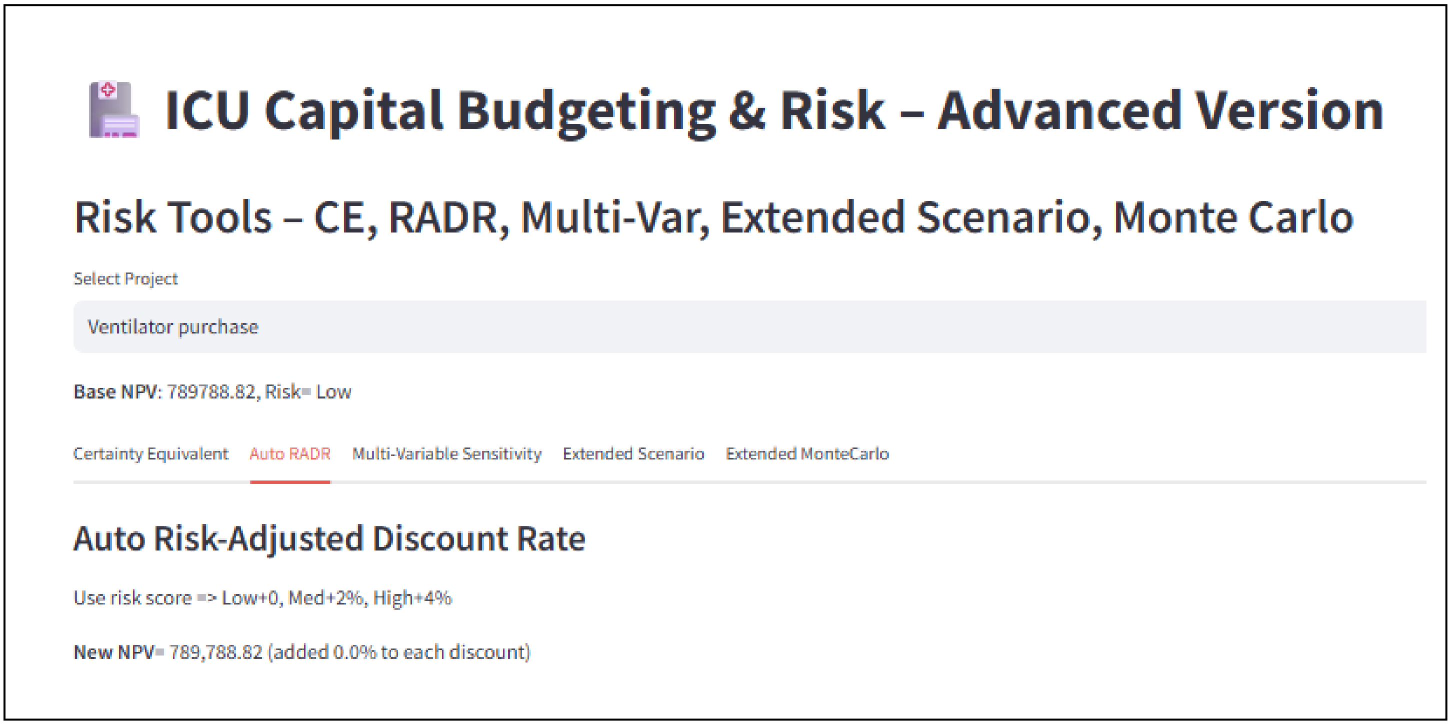

**eFigure 4. Multi-variable sensitivity: revenue –10 %, operating costs +10 %**

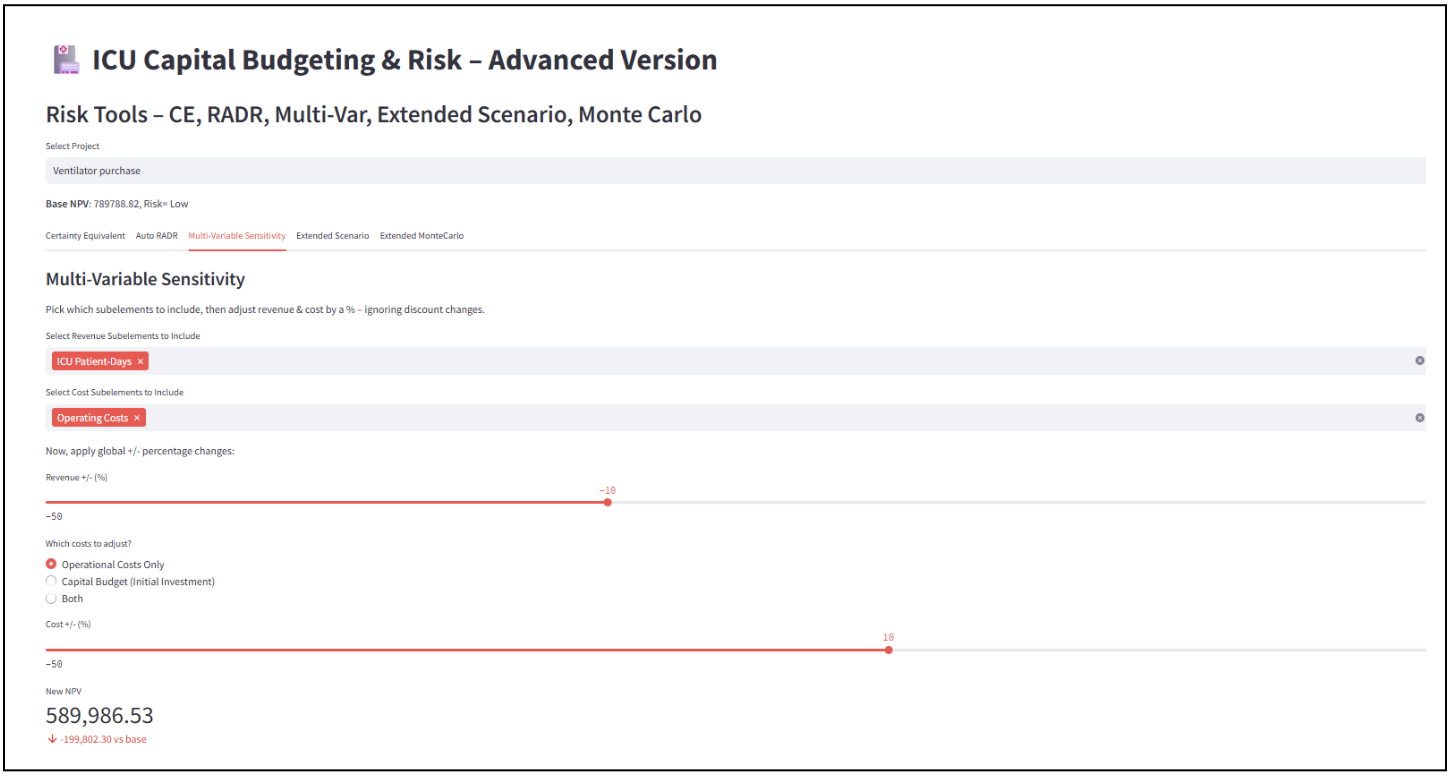

**eFigure 5. Worst-, base-, and best-case NPVs for the ventilator upgrade.**

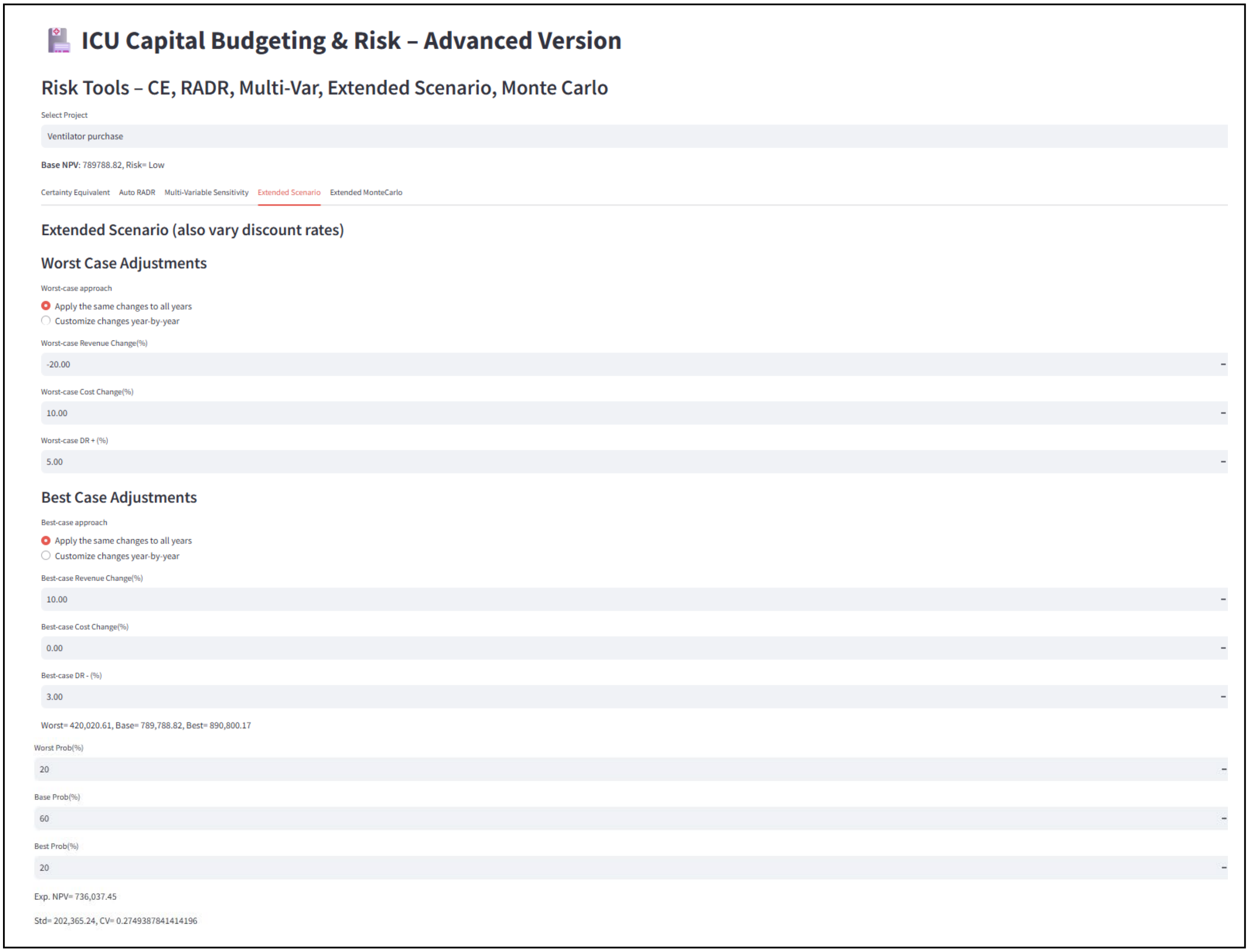

**eFigure 6. Monte-Carlo distribution (500 trials) for ventilator upgrade NPV.**

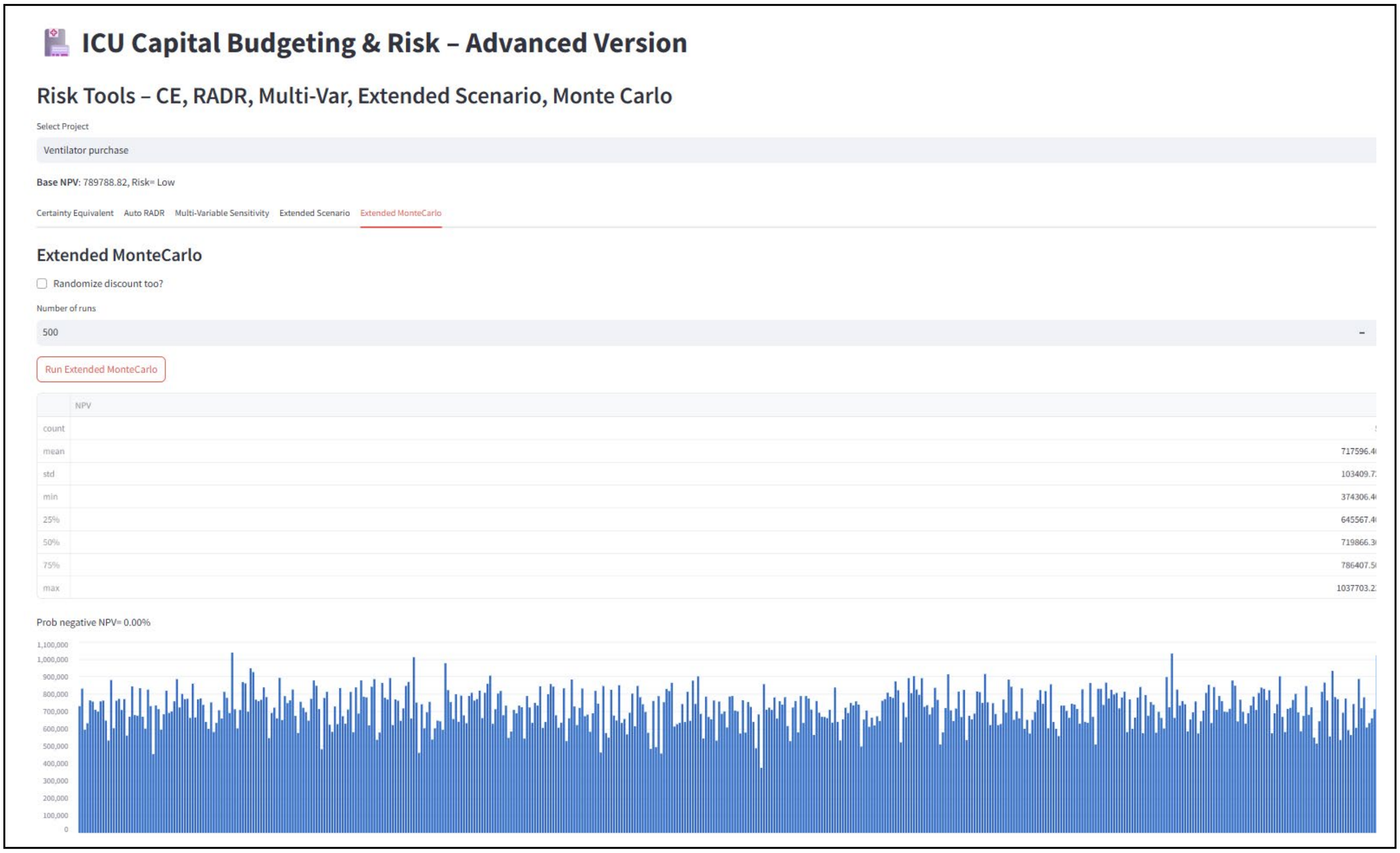

**eFigure 7. GPT-4 multi-year parse for the five-bed ICU expansion.**

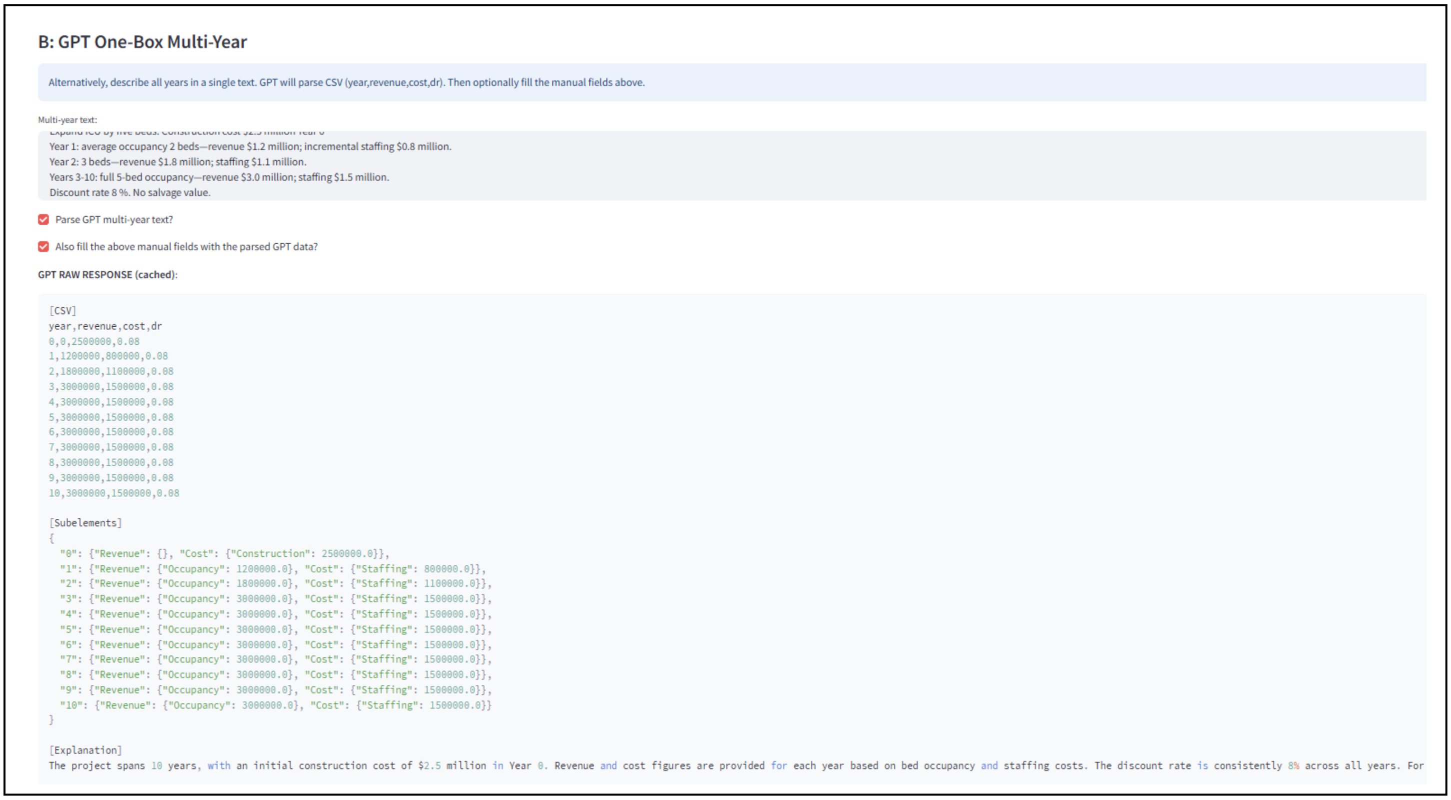

##### B: GPT One-Box Multi-Year

Alternatively, describe all years in a single text. GPT will parse CSV (year,revenue,cost,dr). Then optionally fill the manual fields above.

Multi-year text:

[illegible]
Year 1: average occupancy 2 beds—revenue $1.2 million; incremental staffing $0.8 million.
Year 2: 3 beds—revenue $1.8 million; staffing $1.1 million.
Years 3-10: full 5-bed occupancy—revenue $3.0 million; staffing $1.5 million.
Discount rate 8 %. No salvage value.

☑ Parse GPT multi-year text?

☑ Also fill the above manual fields with the parsed GPT data?

**GPT RAW RESPONSE (cached):**

```
[CSV]
year,revenue,cost,dr
0,0,2500000,0.08
1,1200000,800000,0.08
2,1800000,1100000,0.08
3,3000000,1500000,0.08
4,3000000,1500000,0.08
5,3000000,1500000,0.08
6,3000000,1500000,0.08
7,3000000,1500000,0.08
8,3000000,1500000,0.08
9,3000000,1500000,0.08
10,3000000,1500000,0.08

[Subelements]
{
  "0": {"Revenue": {}, "Cost": {"Construction": 2500000.0}},
  "1": {"Revenue": {"Occupancy": 1200000.0}, "Cost": {"Staffing": 800000.0}},
  "2": {"Revenue": {"Occupancy": 1800000.0}, "Cost": {"Staffing": 1100000.0}},
  "3": {"Revenue": {"Occupancy": 3000000.0}, "Cost": {"Staffing": 1500000.0}},
  "4": {"Revenue": {"Occupancy": 3000000.0}, "Cost": {"Staffing": 1500000.0}},
  "5": {"Revenue": {"Occupancy": 3000000.0}, "Cost": {"Staffing": 1500000.0}},
  "6": {"Revenue": {"Occupancy": 3000000.0}, "Cost": {"Staffing": 1500000.0}},
  "7": {"Revenue": {"Occupancy": 3000000.0}, "Cost": {"Staffing": 1500000.0}},
  "8": {"Revenue": {"Occupancy": 3000000.0}, "Cost": {"Staffing": 1500000.0}},
  "9": {"Revenue": {"Occupancy": 3000000.0}, "Cost": {"Staffing": 1500000.0}},
  "10": {"Revenue": {"Occupancy": 3000000.0}, "Cost": {"Staffing": 1500000.0}}
}

[Explanation]
The project spans 10 years, with an initial construction cost of $2.5 million in Year 0. Revenue and cost figures are provided for each year based on bed occupancy and staffing costs. The discount rate is consistently 8% across all years. For
```

**eFigure 8. Certainty-equivalent analysis for ICU expansion (CE = 0.80).**

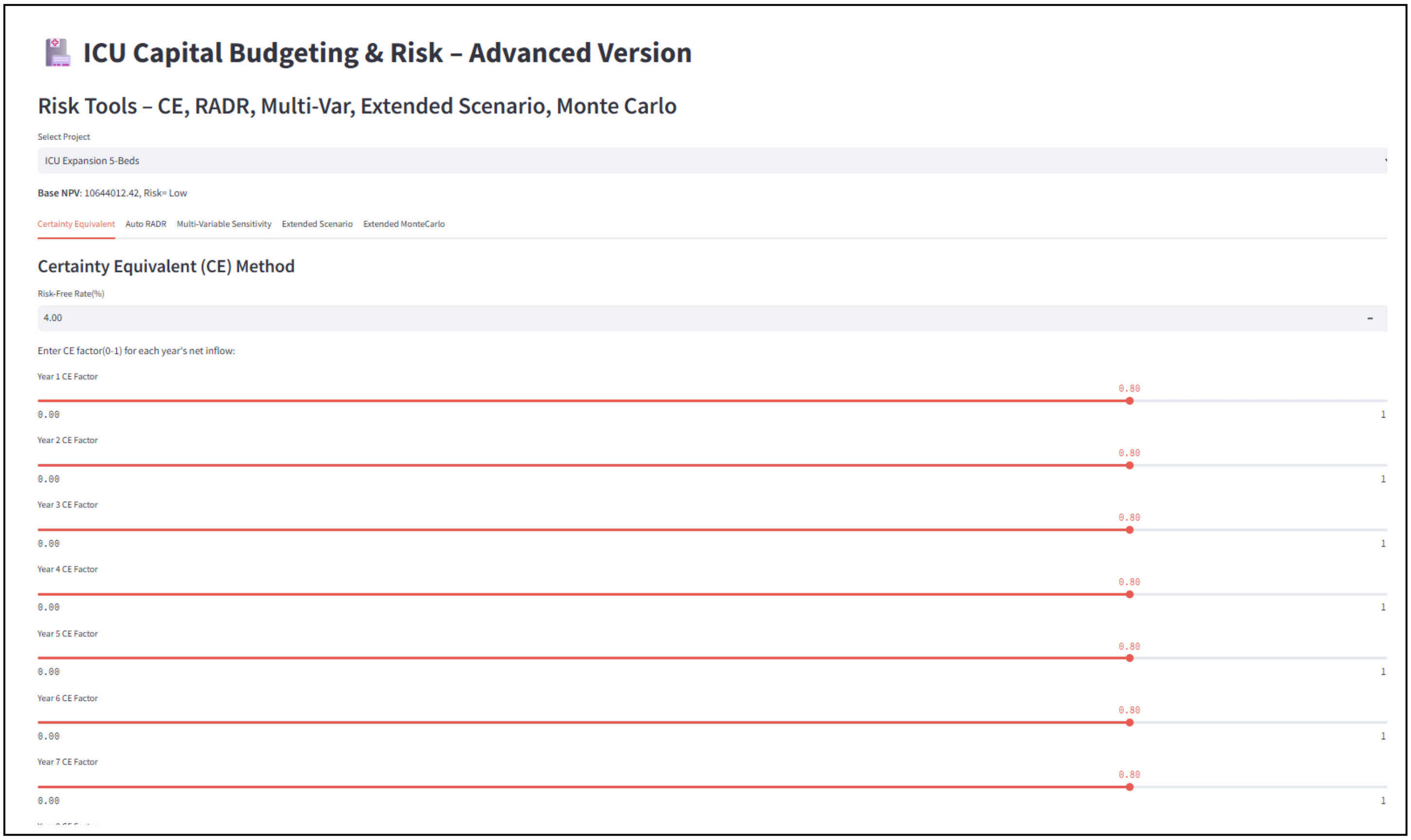

**eFigure 9. RADR output for ICU expansion—Low-risk adjustment (0 pp).**

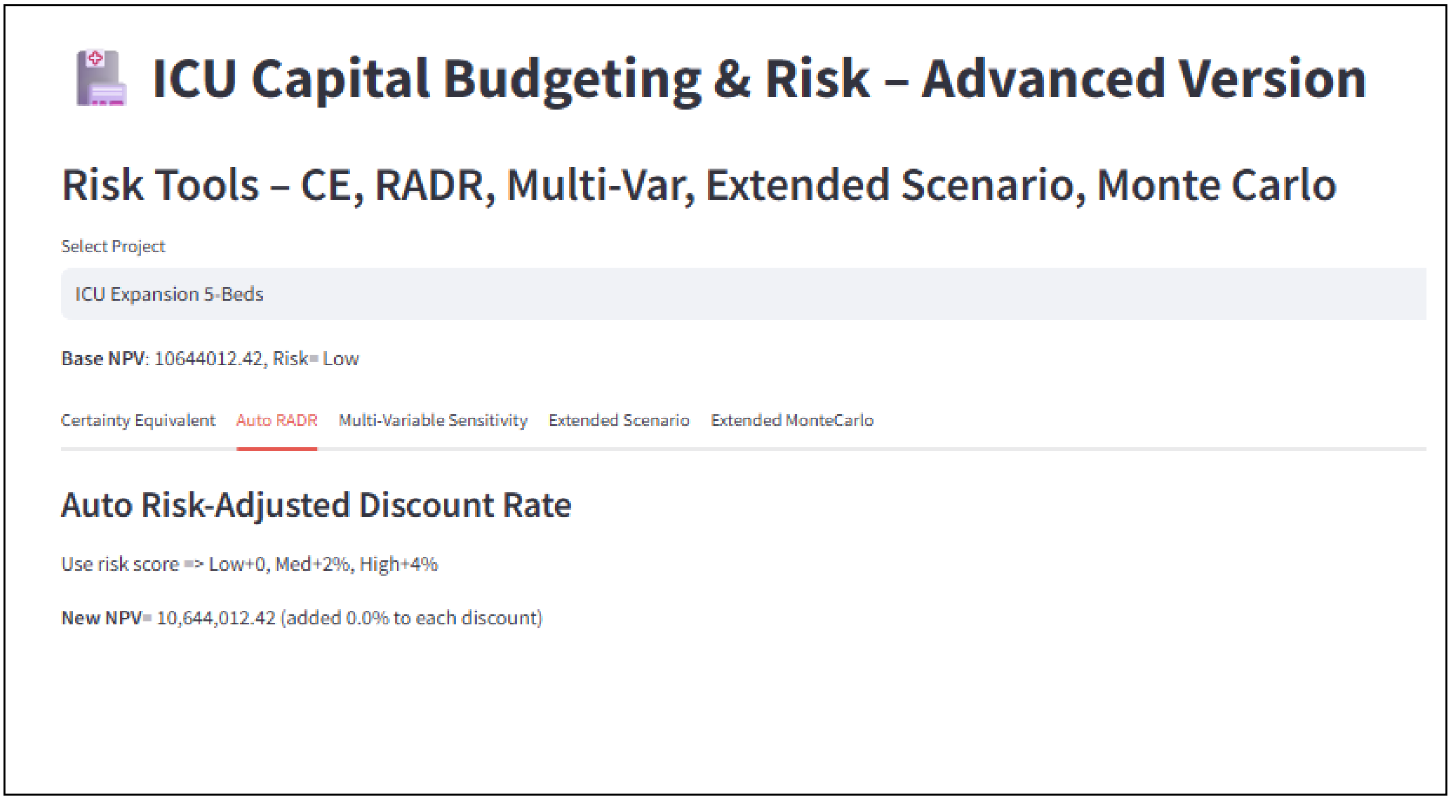

**eFigure 10. Sensitivity: revenue –15 %, staffing cost +10 % (ICU expansion).**

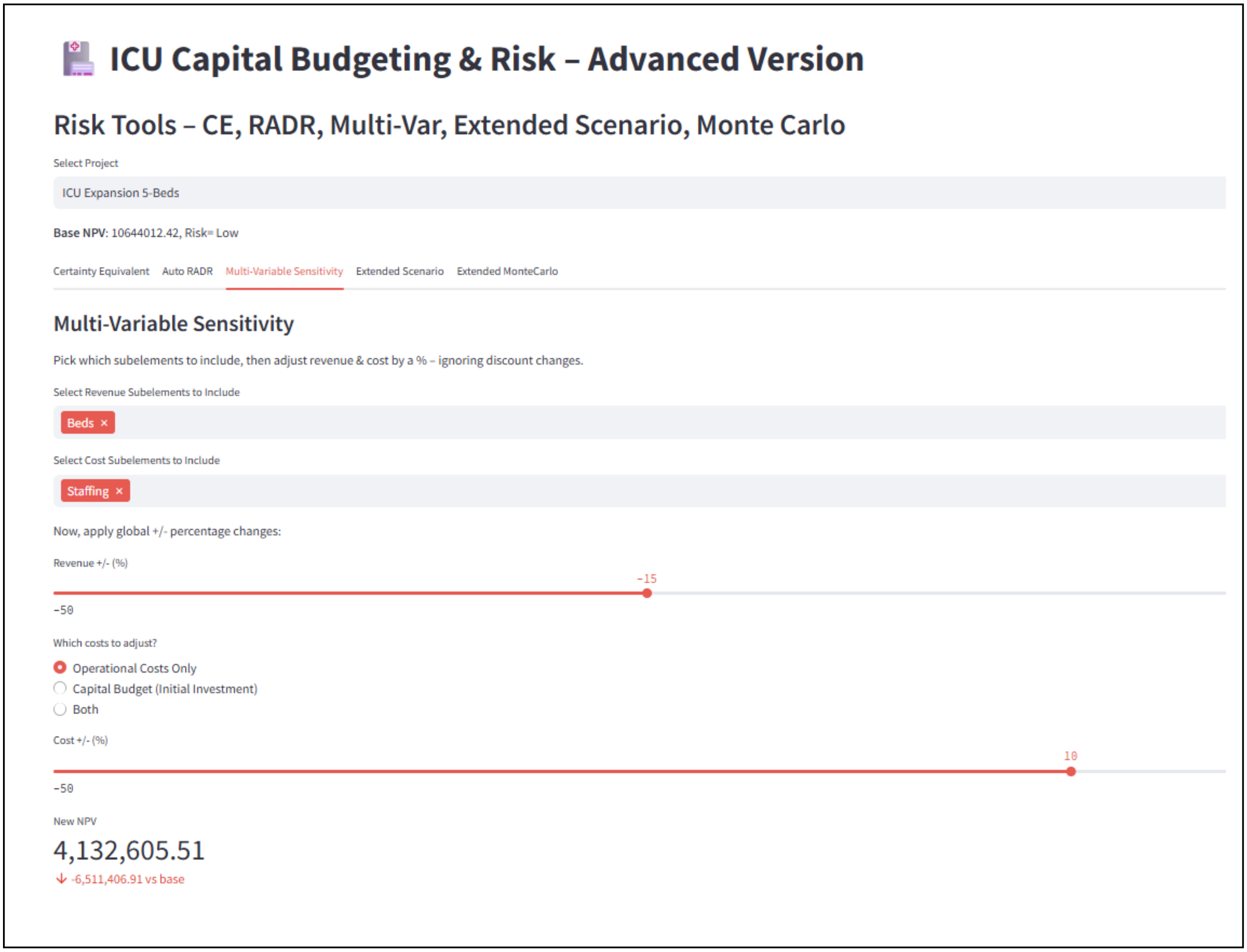

**eFigure 11. Worst/base/best NPVs for ICU expansion (dispersion view).**

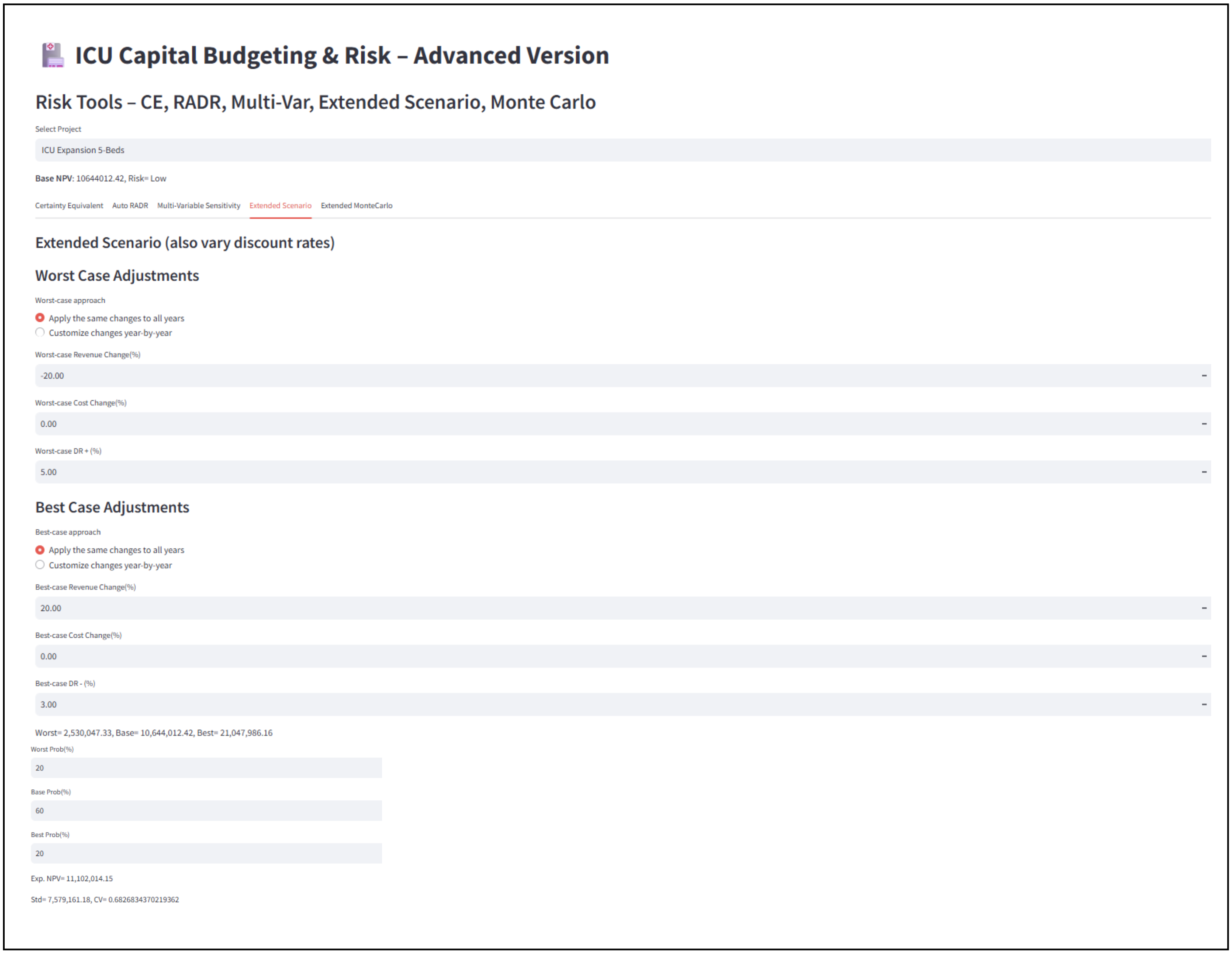

**eFigure 12. Monte-Carlo NPV distribution for ICU expansion (500 trials).**

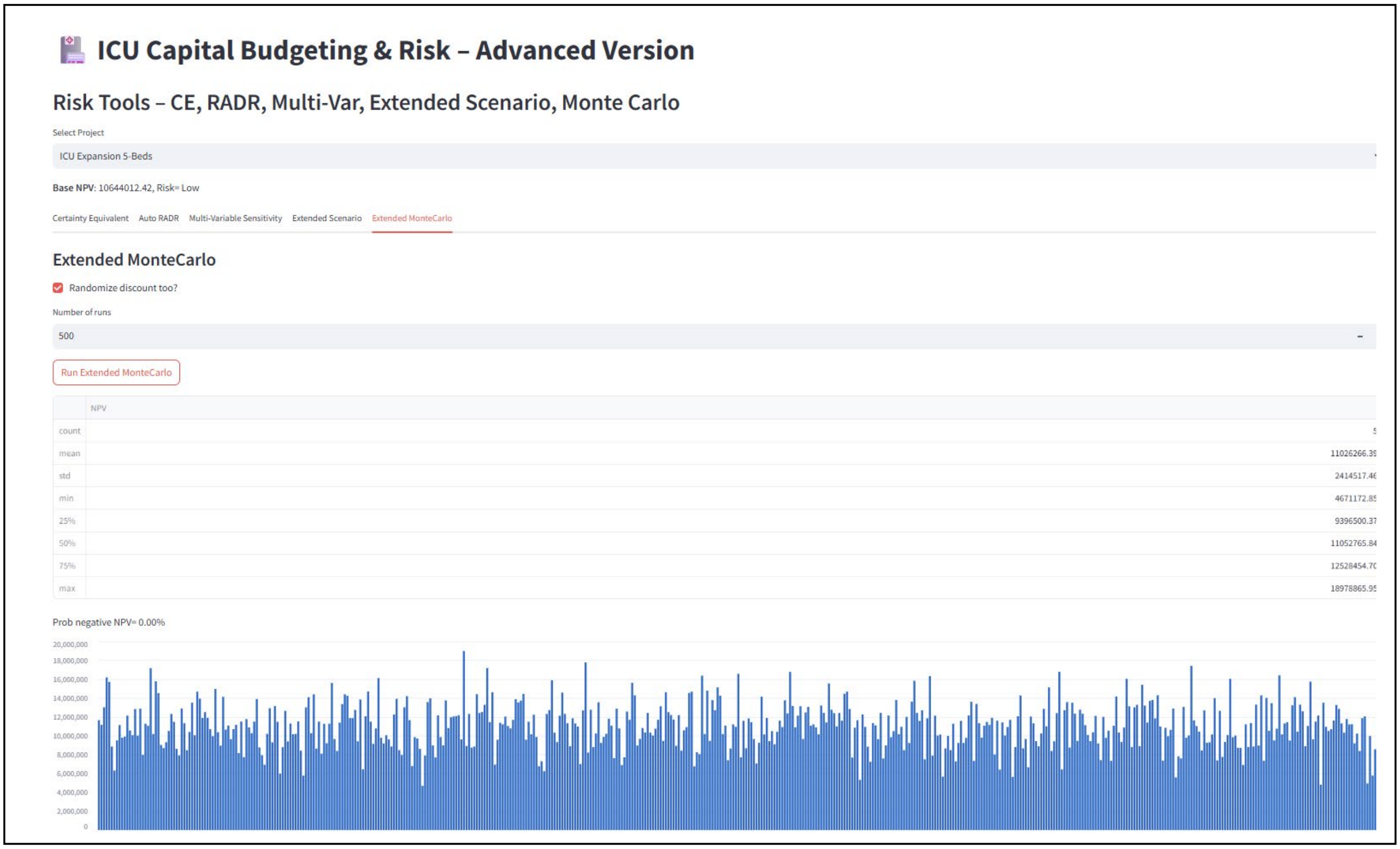

**eFigure 13. GPT-4 multi-year parse for the dialysis cartridge system.**

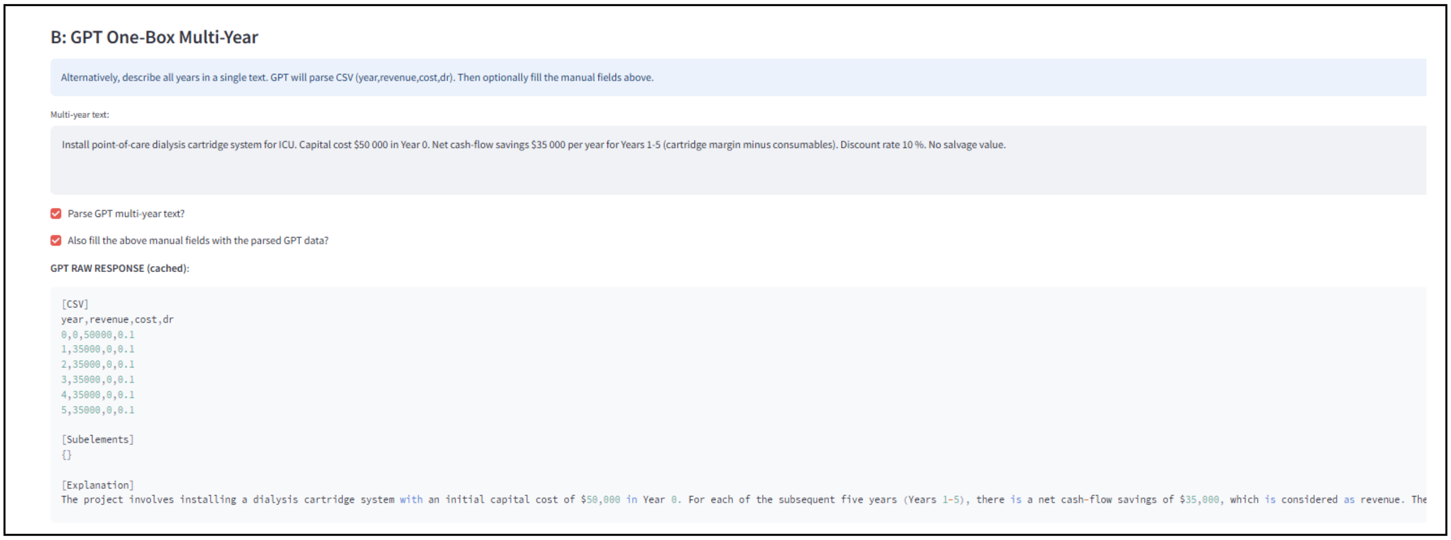

**eFigure 14. Certainty-equivalent analysis for dialysis system (CE = 0.85).**

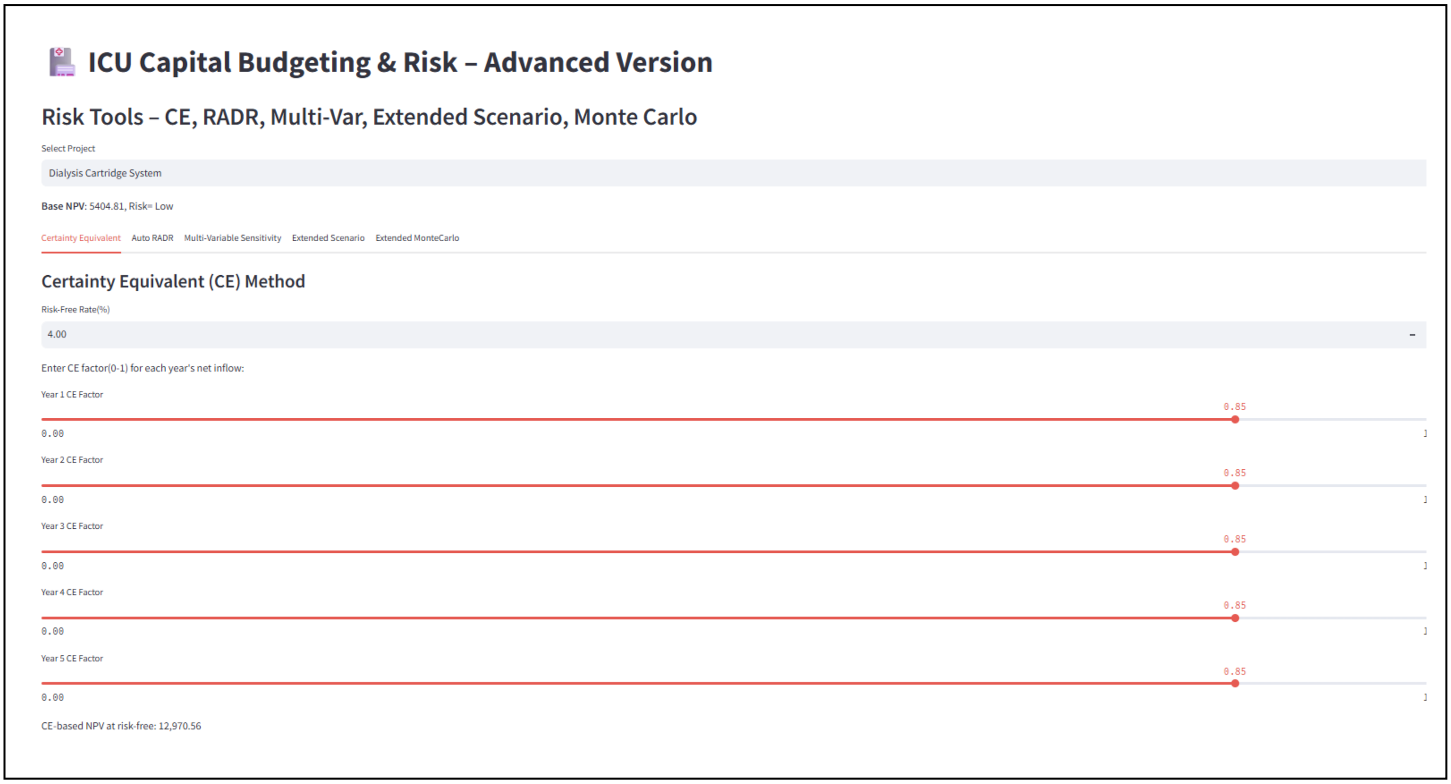

**eFigure 15. RADR output for dialysis system—Low-risk adjustment.**

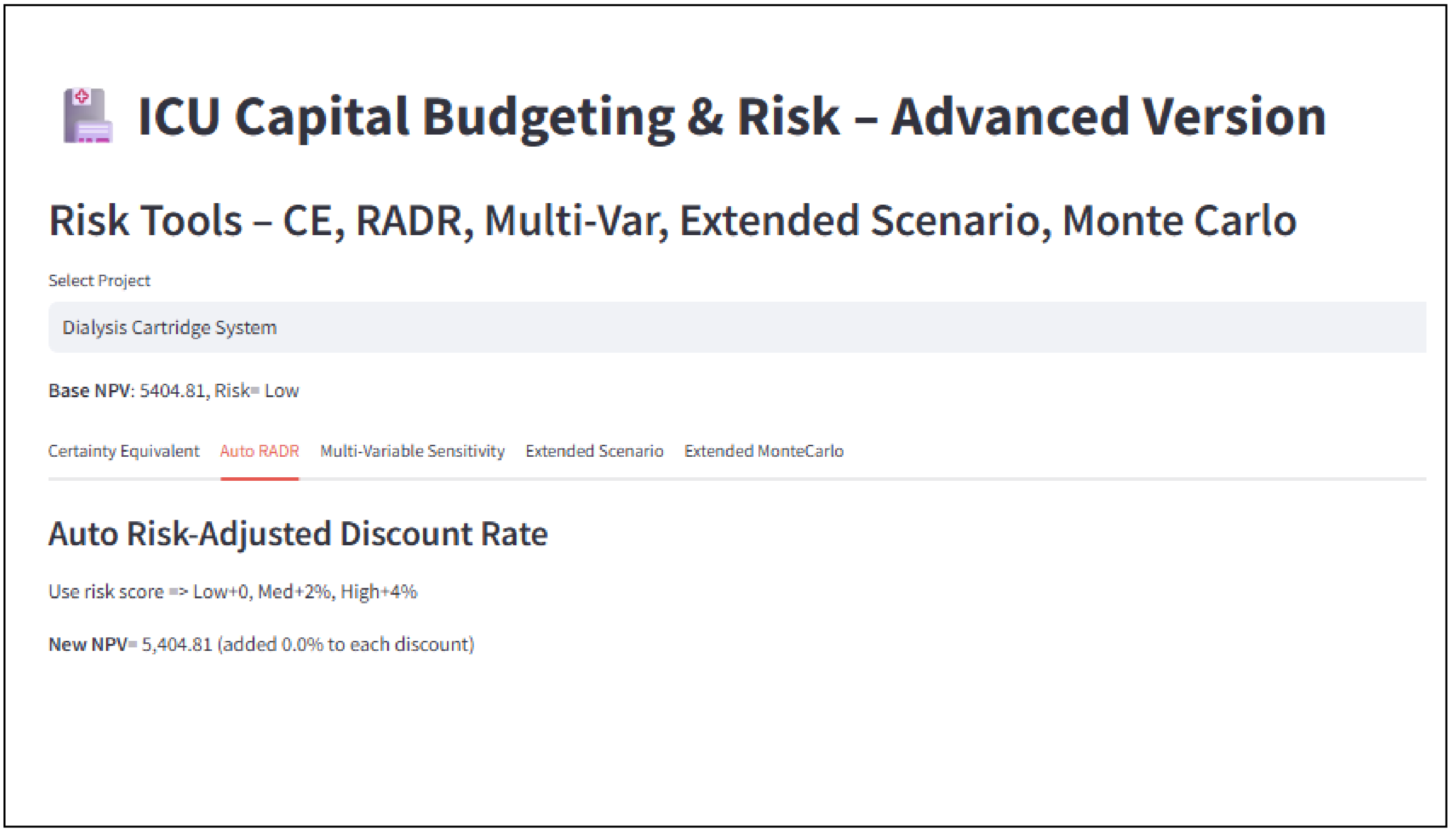

**eFigure 16. Multi-variable sensitivity (no selectable sub-elements) for dialysis system.**

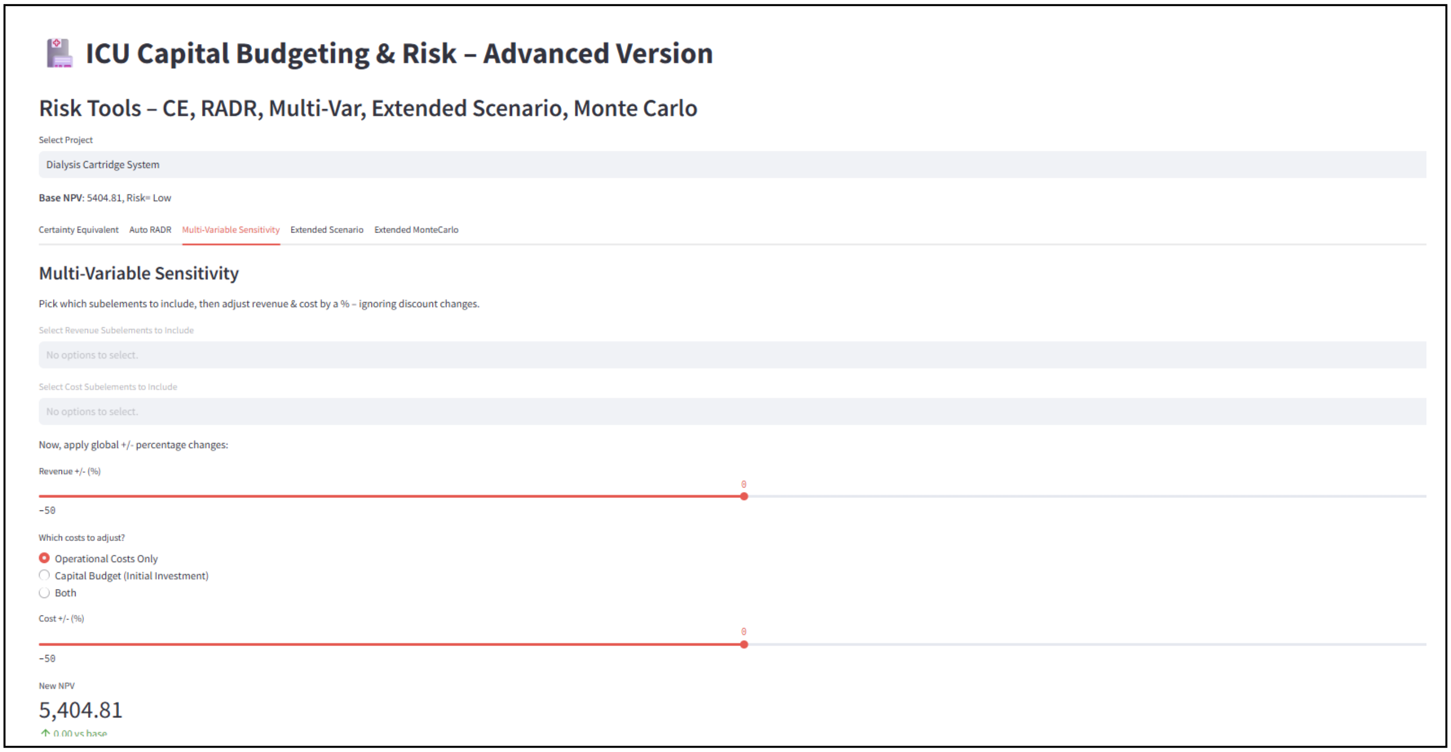

**eFigure 17. Worst/base/best NPVs and expected NPV for dialysis system.**

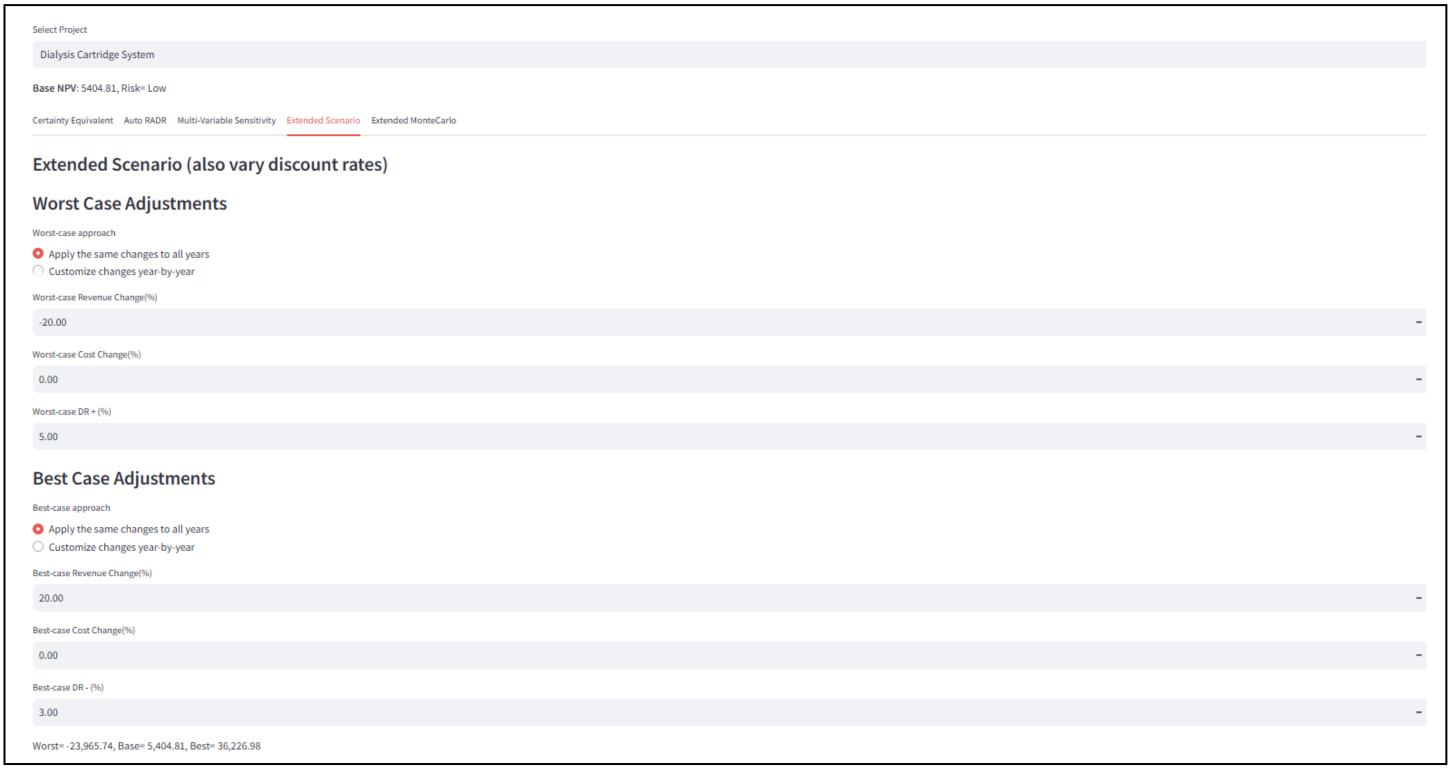

**eFigure 18. Monte-Carlo NPV distribution for dialysis system (500 trials; 34 % negative).**

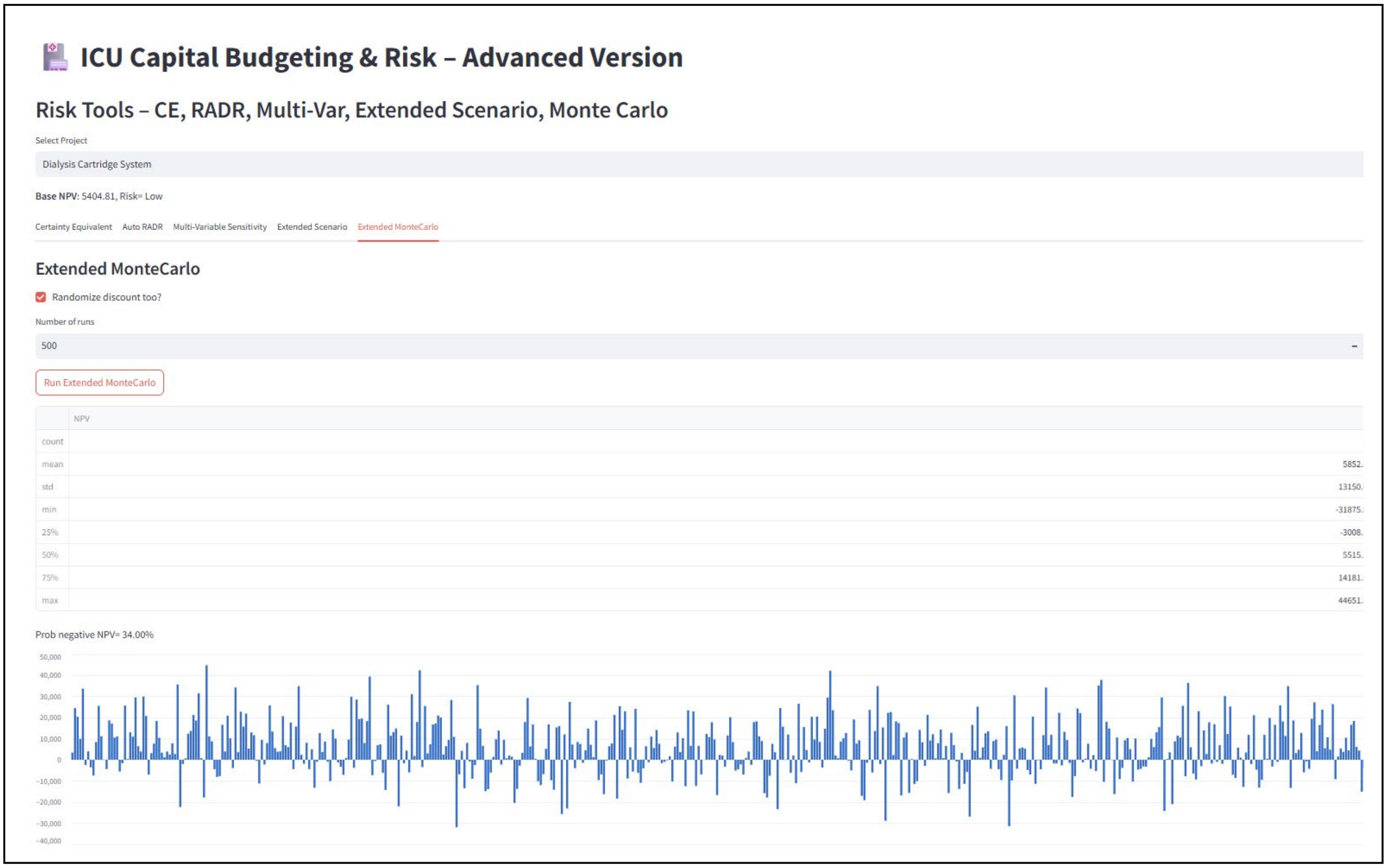

**eFigure 19. Capital-rationing dashboard with expanded $2.67 M budget showing all three projects funded.**

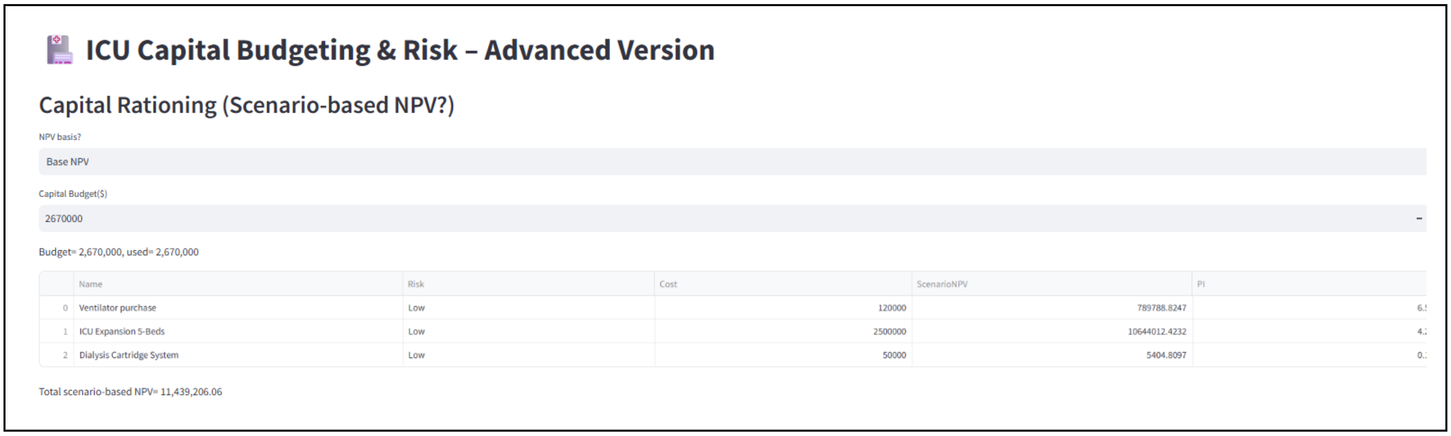

### ICU Capital Budgeting & Risk – Advanced Version

#### Capital Rationing (Scenario-based NPV?)

NPV basis?

Base NPV

Capital Budget($)

2670000

Budget= 2,670,000, used= 2,670,000

| | Name | Risk | Cost | ScenarioNPV | PI |
|---|---|---|---|---|---|
| 0 | Ventilator purchase | Low | 120000 | 789788.8247 | 6.[illegible] |
| 1 | ICU Expansion 5-Beds | Low | 2500000 | 10644012.4232 | 4.[illegible] |
| 2 | Dialysis Cartridge System | Low | 50000 | 5404.8097 | 0.[illegible] |

Total scenario-based NPV= 11,439,206.06
